# Mobilefuge: A low-cost, portable, open source, 3D-printed centrifuge that can be used for purification of saliva samples for SARS-CoV2 detection

**DOI:** 10.1101/2021.01.06.21249280

**Authors:** Mohini Bhupathi, Ganga Chinna Rao Devarapu

## Abstract

One of the best ways to contain the spread of COVID-19 is frequent testing of as many people as possible and timely isolation of uninfected personnel from infected personnel. However, the cost of massive testing is affordable in many countries. The existing technologies might not be scalable to offer affordable testing for millions of people. To address this issue, novel testing methods based on Loop-Mediated Isothermal Amplification (LAMP) were proposed that are more sensitive, require less reagents and can work with saliva samples instead of more tedious nasal swabs. As a result, LAMP based protocols can make it possible to drive the cost down to one dollar per test. These LAMP based methods require a centrifuge device, mostly for separation of viral particles from reaction inhibitors in saliva samples. However, centrifuge is neither accessible nor affordable in many resource limited settings, especially during this pandemic situation when normal supply chains are heavily disrupted. To overcome these challenges, we invented a low-cost centrifuge that can be useful for carrying out low-cost LAMP based detection of SARS-Cov2 virus in saliva. The 3D printed centrifuge (Mobilefuge) is portable, robust, stable, safe, easy to build and operate. The Mobilefuge doesn’t require soldering or programming skills and can be built without any specialised equipment, yet practical enough for high throughput use. More importantly, Mobilefuge can be powered from widely available USB ports, including mobile phones and associated power supplies. This allows the Mobilefuge to be used even in off-grid and resource limited settings. We believe that our invention will aid the efforts to contain the spread of COVID-19 by lowering the costs of testing equipment. Apart from the COVID-19 testing, the Mobilefuge can have applications in the field of biomedical research and diagnostics.

## Introduction

The current pandemic, COVID-19 is spreading fast across the globe resulting in loss of millions of lives [1] and livelihoods [2]. Many countries are now witnessing second and third waves with newly mutated SARS-CoV2 looking to create further havoc. One of the effective ways to decrease these devastating effects of the COVID-19 is to carry out as many tests as possible and as frequently as possible. It is also important to obtain the results of these tests available promptly, so that infected cases can be isolated for treatment while the uninfected can be protected.

The current SARS-CoV2 testing methods are broadly based on four techniques: Polymerase Chain Reaction (PCR), Loop-mediated isothermal amplification (LAMP), Lateral flow and Enzyme-Linked Immunosorbent Assay (ELISA) [3], [4]. Out of these four techniques, LAMP has high potential to be scaled up for point-of-care, near-patient-care and point-of-entry applications as it is highly reliable, sensitive, faster, cost-effective and doesn’t require complex equipment [5], [6], [7]. Due to these benefits, a number of LAMP assays have been fast tracked by FDA for first at-home tests for COVID-19 [4,8] [9], [10] and have the potential to bring down the cost of each test to less than a dollar [11–14].

In particular, a LAMP assay that was developed at Harvard University can detect single virtual RNA per microliter from saliva samples [12]. This is significant as it eliminates current elaborate sample collection methods through nasal swabs, thereby decreasing the professional required and associated risks and further reducing the costs and resources. However, this Harvard assay and similar low-cost LAMP [11] assays require a centrifuge to separate viral RNA in saliva from other inhibitors.

Without the centrifugation step, saliva based LAMP tests are less sensitive [14], [15] down to fifty viral particles per microliter from one viral particle per microliter [12]. As a result, LAMP assays would be limited only to identify individuals with moderate or high viral load and would not be suitable to detect viral infections in individuals with low viral load such as individuals with onset of infection or recovering from the infection.

Furthermore, a recent assay that combines Recombinase Polymerase Amplification and LAMP was proposed to achieve high sensitivity and selectivity [12], [16]. Without a centrifuge such LAMP assays require 65% more reagents which will lead to an increase of cost per test. Therefore, centrifuge is a crucial equipment required for LAMP assays for separating the viral RNA in saliva from other inhibitors to increase the wide adoption of LAMP assays to achieve affordable SARS-CoV2 testing.

However, due to the pandemic situation that disrupted the global supply chains [17], medical equipment and reagents are still in short supply across the globe [18,19]. Furthermore, many resource limited regions with constrained medical facilities might not afford commercially available medical equipment [20], [21]. To alleviate this issue, recently Prakash’s lab from Stanford University demonstrated a low-cost centrifuge that was realised by cleverly exploiting a hand crank rotation mechanism in LED torch [11] for a low-cost LAMP assay. There are many other similar low-cost centrifuges in the literature that could be useful for this purpose and we believe Mobilefuge will be a promising inclusion to the list.

Centrifuge is a simple and intuitive scientific device as it involves a single rotary mechanism, something that mankind has been exploiting since the invention of the wheel. Therefore, scientists and hobbyists exploited a variety of existing rotary mechanisms in consumer devices and toys to build centrifuges. These highly creative designs involve either a hand powered rotary mechanism in hand crank torch lights [11], fidget spinners [22], [23], toys [24], [25], salad spinners [26], [27], egg beaters [28] or electric motor powered rotary mechanism in consumer electronic devices such as drones [29], [30], [31], [32], [33], [34] [35], [36], computer fans [37], hard disks [38], DVD drives [39] and Dremel tools [40]. Proliferation of low-cost digital fabrication methods such as 3D printers [22], [40], CNC machines [33] and laser cutters [11], [32], [38] helped to bring the best out of these designs by providing a safety enclosure and customisable rotor for various centrifuge tubes.

All the above mentioned centrifuge designs serve different purposes varying from pedagogic demonstrations [26], [32], [40] to medical diagnostics [22], [23]. Hand powered centrifuges might not be practical for high throughput use in medical or scientific labs and offer limited safety while in operation. On the other hand, most of the centrifuge designs that involve electric motors require high power and complex fabrication techniques. This is understandable as they were designed for general lab purposes which require high speed with speed control options, thus requiring many components and complex build procedures.

However, the current pandemic situation poses a different challenge that requires a simple centrifuge design that does not need to have high rotational speed, yet practical enough to be deployed for diagnostic purposes. To address this challenge, we designed a 3D-printed centrifuge henceforth referred to as Mobilefuge. The Mobilefuge consists of a USB fan motor that can be powered by a USB power source.

The main objective of the Mobilefuge is to build a stable, scalable, portable, affordable, robust, and easy to build centrifuge that can be powered using widely available 5V USB outlets, including smartphones. Most of the smartphones supply 5V from their charging ports as part of the USB On-The-Go specification standard [41]. Therefore, the ability to run the Mobilefuge directly from smartphones makes it possible to deploy the Mobilefuge on-grid and off-grid settings. Currently, there are estimated 3.5 billion smartphones [38] in the world making smartphones as the most widely accessible portable electric device across the globe. Along with smartphones, other mobile phones, and their accessories such as chargers and rechargeable battery banks provide 5V output. As a result, 5V power outlet is the most widely used power source which Mobilefuge leverages to reach resource limited regions. This is highly desirable particularly during this pandemic situation as the Mobilefuge can be taken to remote locations and employed for both near-patient and point-of-care diagnostic purposes.

In the following, we will describe the fabrication of Mobilefuge and evaluate its performance to generate the speeds required for LAMP assays. Finally, we discuss the benefits of Mobilefuge such as portability, safety, and scalability in comparison with the other existing centrifuge designs for resource limited settings in the fight against COVID-19.

## Materials and Methods

The Mobilefuge mainly consists of four parts as shown in Fig. 1: 3D-printed enclosure and Rotor 2) USB fan motor 3) Female to male USB cable 4) Self-adhesive damping pad (Optional).

**Figure 1:**
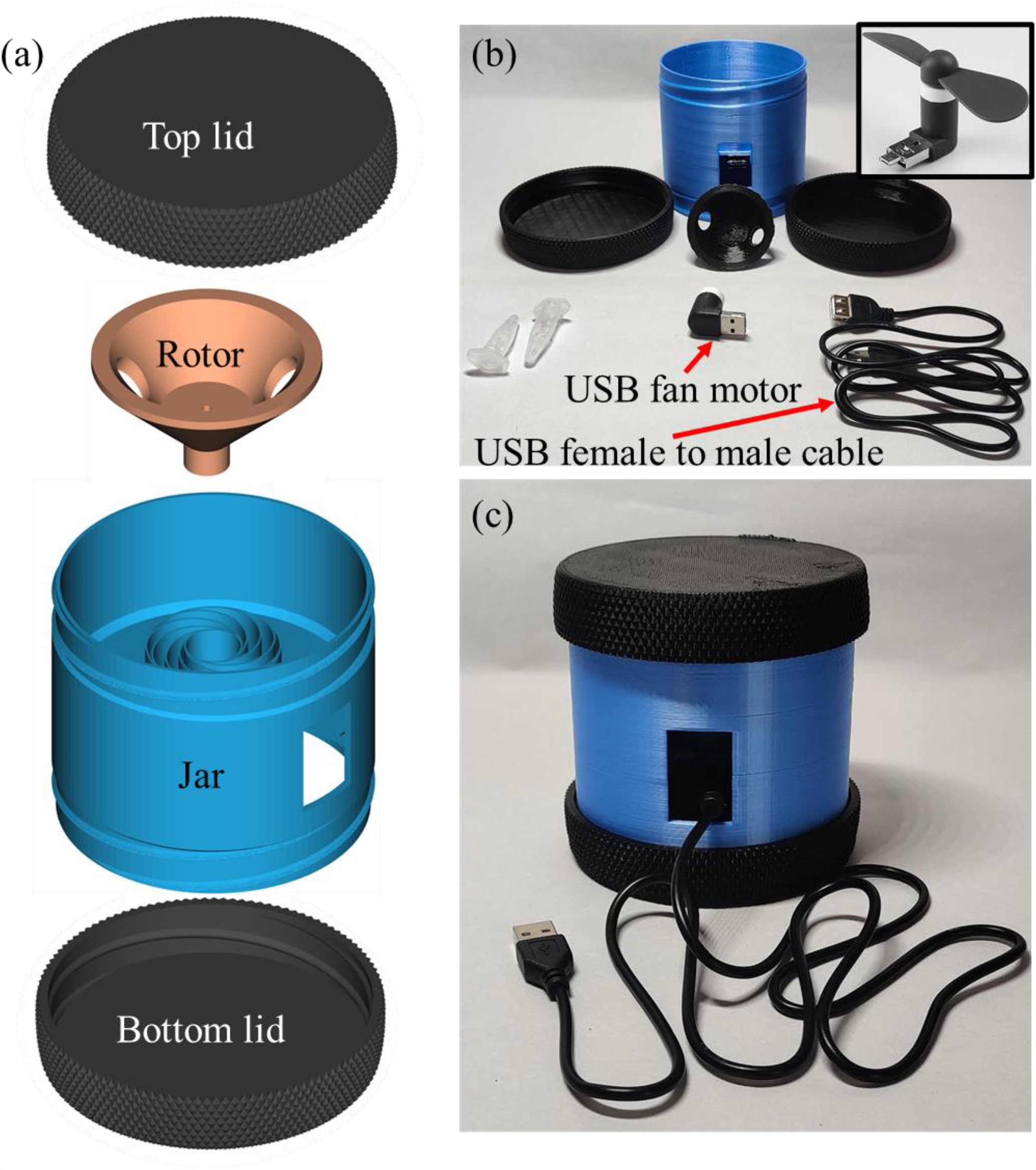
(a) CAD diagrams of 3D printed parts required for the Mobilefuge (b) The 3D printed parts and the electrical parts required to fabricate the Mobilefuge. The USB fan that powers the Mobilefuge is shown in the inset (c) Completely assembled Mobilefuge.

The enclosure for the Mobilefuge consists of three parts as shown in Fig. 1(a): top lid, main jar, and bottom lid. The top and bottom lids are locked to the main jar through the threads which makes the Mobilefuge’s assembly easy. The main jar consists of a spiral placeholder to hold the USB fan motor in place. The spiral placeholder helps in stabilising the motor vibrations [33]. The bottom lid provides room for the USB plug and the top lid provides safety while the centrifuge is in operation. Along with these enclosure parts, a rotor is designed to press-fit to the motor axis and hold two centrifuge tubes.

All the above mentioned CAD parts are designed using open-source parametric CAD software OpenScad [42]. The design of the enclosure, spiral placeholder and the rotor are adopted from various open source 3D CAD designs in Refs. [31], [43], [44] and respectively. These 3D-designs are then exported as STL files and translated into a list of instructions to the 3D-printer, *i*.*e*., G Code by using a CURA software which slices the 3D model into 2D layers. All the Openscad and STL design files of the Mobilefuge are made freely available [45]. Finally, these designs are printed using a Monoprice select Mini V2 3D-printer [46] using PLA material with following settings: 10% infill and support structure. Support structures are removed after the objects are 3D printed.

Other than the above 3D-printed parts, a USB fan motor is used to power the Mobilefuge (See inset in Fig. 1(b)). These USB fans are ubiquitously available as consumer devices. They contain a USB/microUSB male connector and can be plugged directly into USB/microUSB ports on laptops, smartphones, and power banks. The USB fans have speeds in the order of 7000-10000 Rotations per minute (RPM) to circulate the air, therefore highly suitable for the centrifuge applications. Once this motor is fixed inside a Mobilefuge jar enclosure, a USB female to male cable is used to connect the fan to a USB power source as shown in Fig. 1(b) and (c). Instead of this female to male USB connector, one can use an OTG USB cable to power the Mobilefuge using a smartphone. All these 3D-printed and electrical parts are assembled as shown and described in Fig. 2 and Video V1 to realise the Mobilefuge.

**Figure 2:**
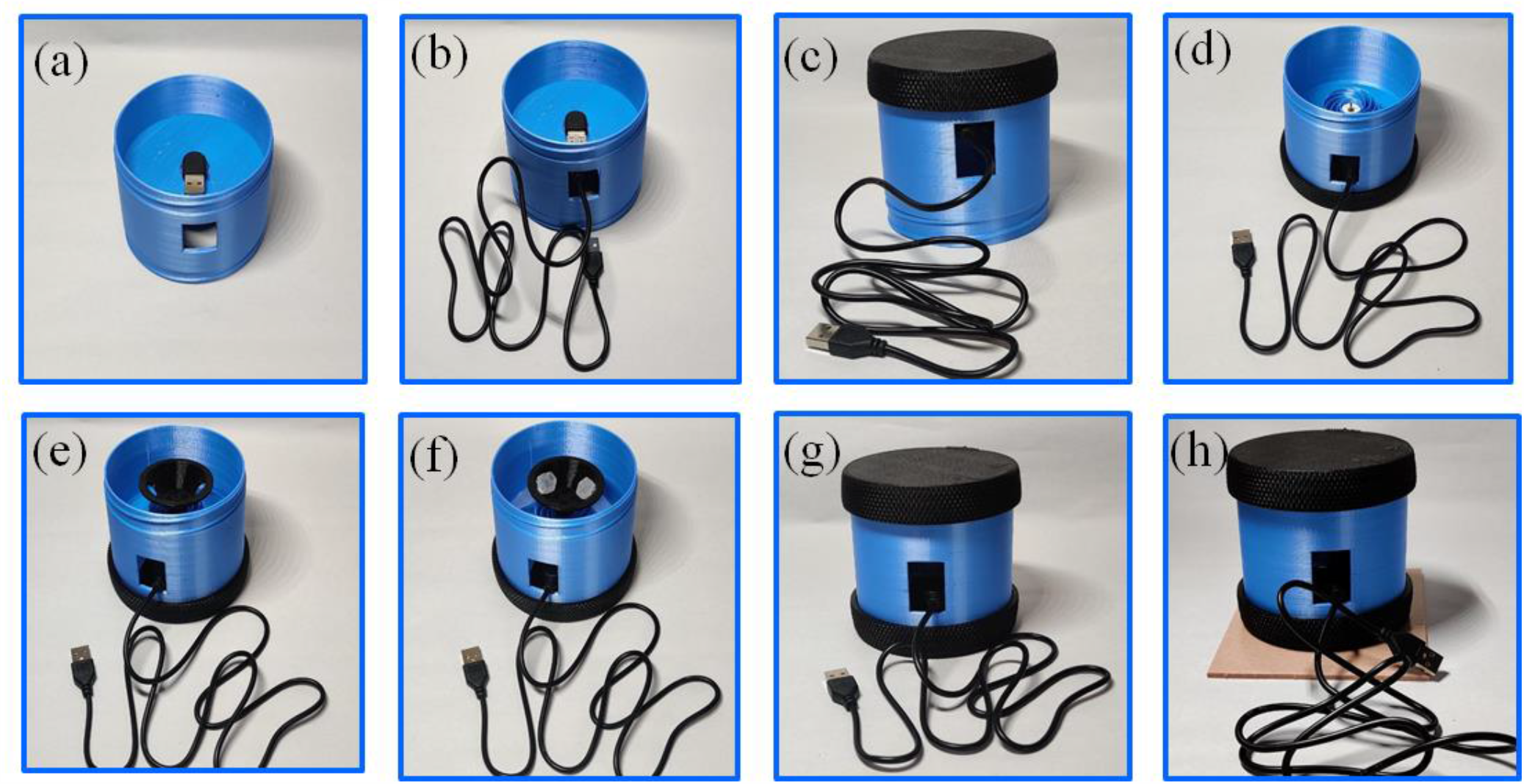
Assembly instructions of the Mobilefuge: (a) Fan motor is press-fitted at the bottom of the jar (b) A USB cable is connected to the motor (c) Bottom lid is fixed to the jar part of the enclosure. (d) Same as (c) but showing the top view of the jar with the bottom lid in place (e) Rotor is press-fitted to the fan motor (f) Centrifuge tubes inserted in the rotor (g) Top lid is threaded to the main jar of the enclosure to complete the assembly of the Mobilefuge. (h) A self-adhesive damping pad is placed at the bottom of the Mobilefuge to improve the stability of Mobilefuge.

### Calculation centrifugal force

The performance of a centrifuge is measured in the units of Relative centrifugal field (RCF). The RCF describes and compares the strength of the acceleration applied by the centrifuge on a sample relative to the earth’s gravitational force.

Centrifugal field on a particle with a mass *m* is given by *F*_*c*_=*mω*^2^*R* where *R* is the distance between the centrifuge tube and the center of the rotor (See Fig. 3 (a)) and *ω* is the rotational speed of the centrifuge. The earth’s gravitational force on the same particle is *F*_*g*_*=mg* where *g* is the earth’s gravitational constant.

**Figure 3:**
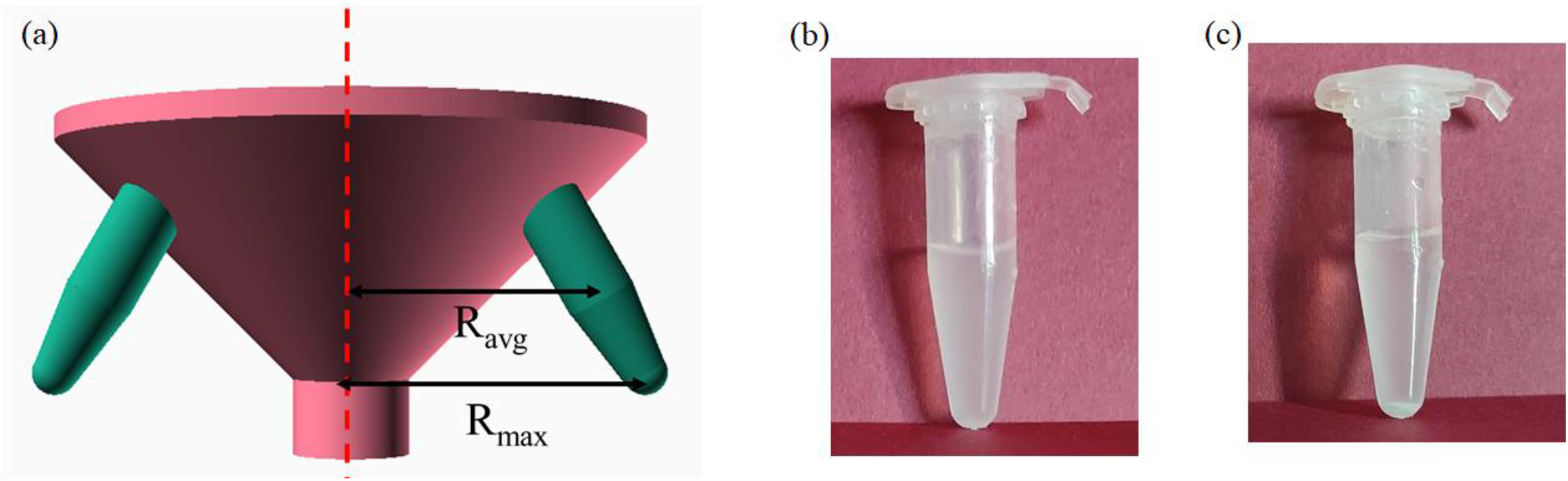
(a) Schematics to show the distance “R” between the rotor and a centrifuge tube used in Eq. (2). (b) “Glass milk” solution with silica particles in a 2mL centrifuge before the centrifugation (c) Pellet of silica particles formed after centrifugation of the “glass milk” solution with Mobilefuge for 30 seconds.

The Relative Centrifugal Field (RCF) can be expressed mathematically as [47,48]:

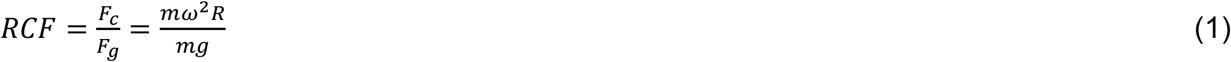

By substituting, *g*=9.8*ms*^−2^and 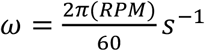, the above equation yields:

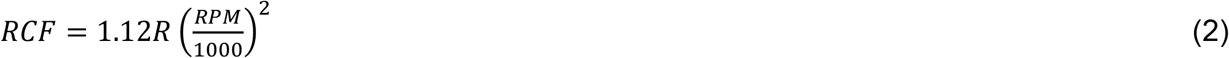

From the above equation, RCF can be measured given the rotational speed (RPM) and the distance (R) from centre of the rotor to the centrifuge tube values of a centrifuge in millimetres. We measured the rotational speed of the Mobilefuge using a non-contact Tachometer (AT6, Tenmacorp) and R values using a Vernier Callipers device.

We found that motor spins at the speed of ∼7500 RPM. The corresponding average and maximum RCF of the Mobilefuge can be estimated from Eq. (2) as 1890 and 2835 for R_avg_ = 30 mm and R_max_ = 45 mm (see Fig. 3(a)) respectively. These rotational speeds and RCFs are more than sufficient to perform the LAMP assay for detection of SARS-Cov2 as described in Refs. [11,12].

### Preparation of “Glass milk” solution

To demonstrate that Mobilefuge can be useful for the purification process during the LAMP assays, we separate the silica particles from a suspension consisting of water and “glassmilk” using the Mobilefuge. The “glass milk” suspension along with Chaotropic salts acts as a binding mix in the low-cost LAMP assays to capture the viral RNA from the cleared saliva solution containing the viral particles [11], [12]. This “glass milk” solution suspension was the predecessor to the commercial silica-based column purification kits. The “glass milk” solution is usually prepared by mixing equal amounts of distil water and silica particles after removing the fine silica particles [49], [50].

So, we prepared the “glass milk” suspension by adding 5 grams of 325 mesh silicon dioxide (sigma 342890) to a tube consisting of 50 ml of water. We waited until the silica particles were precipitated at the bottom of the tube and then the supernatant was poured off to remove the fine silica particles smaller than 1 micron from the solution. Then another 50 ml water is added to the tube with the precipitated silica particles to make the “glass milk” suspension. The centrifugation outcome of this “glass milk” suspension with the Mobilefuge is described in the Results section.

## Results

### Evaluating performance of the Mobilefuge

As explained in the earlier section, we will demonstrate the centrifugation ability of the Mobilefuge by separating the silica particles from a suspension consisting of water and “glass milk”. This centrifugation process mimics the sample purification step in the LAMP assay protocols where viral RNA separated from the cleared saliva solution. For this purpose, 5 uL of “glass milk” is added to 500 uL of distilled water in a 2mL centrifuge tube. This 2 mL centrifuge tube is placed in the Mobilefuge and spun for 30 seconds. We show the image of the centrifuge tube before and after the centrifugation in Fig. 3(b) and (c) respectively. It is clear from these images that silica particles are trapped at the end of the centrifuge tube as a pellet while leaving the water as a supernatant. In LAMP assays, the nucleic acids of the SARS-CoV2 virus particles present in the saliva will bind to the silica particles and become a pellet at the end of the centrifuge tube. After pouring off the supernatant, this pellet containing the viral nucleic acids is retained for further processes in a LAMP assay. Note that LAMP assay for detection of SARS-CoV2 is not carried out in this article.

### Evaluating stability of the Mobilefuge

Equipment such as a centrifuge that consists of rotating parts is prone to vibrations due to unbalanced rotors which will lead to noise. In the case of Mobilefuge, since the rotor is 3D-printed, it is sometimes difficult to mount it exactly at the center of the motor. Similarly, any small difference in the weights of the centrifuge tubes will also lead to an unbalanced rotor and consequently will result in vibrations. It is highly desirable to have a stable operation of centrifuge for long term durability and safety purposes.

Therefore, we measured the stability of Mobilefuge by indirectly measuring the vibrations on a surface where the Mobilefuge is placed. In particular, we measured the vibrations caused by the Mobilefuge loaded with two 2 mL centrifuge tubes each with 1mL of water on a table. For this purpose, we recorded data from inbuilt accelerometers in a smartphone (Samsung Galaxy note 10) placed on the same table surface using Phyphox smartphone application [51]. Phyphox is an open-source smartphone application built to perform experiments using inbuilt sensors in smartphones. Along with this accelerometer data, the noise generated while the Mobilefuge is in operation is also recorded with the microphone in the same smartphone and Phyphox smartphone application.

In Fig. 4, we show the accelerometer sensor data (sampled every 20 ms) from the smartphone plotted against the time. The left (0-10 s) and the middle (10-20 s) panels represent the accelerometer data when the Mobilefuge is not in use and in use respectively. The red, green and blue lines indicate the accelerometer data in x, y and z directions respectively. The corresponding standard deviations “σ” of this accelerometer data are indicated above these lines. It is evident from the accelerometer data and standard deviation values that Mobilefuge causes considerable vibrations on the table surface when in use.

**Figure 4:**
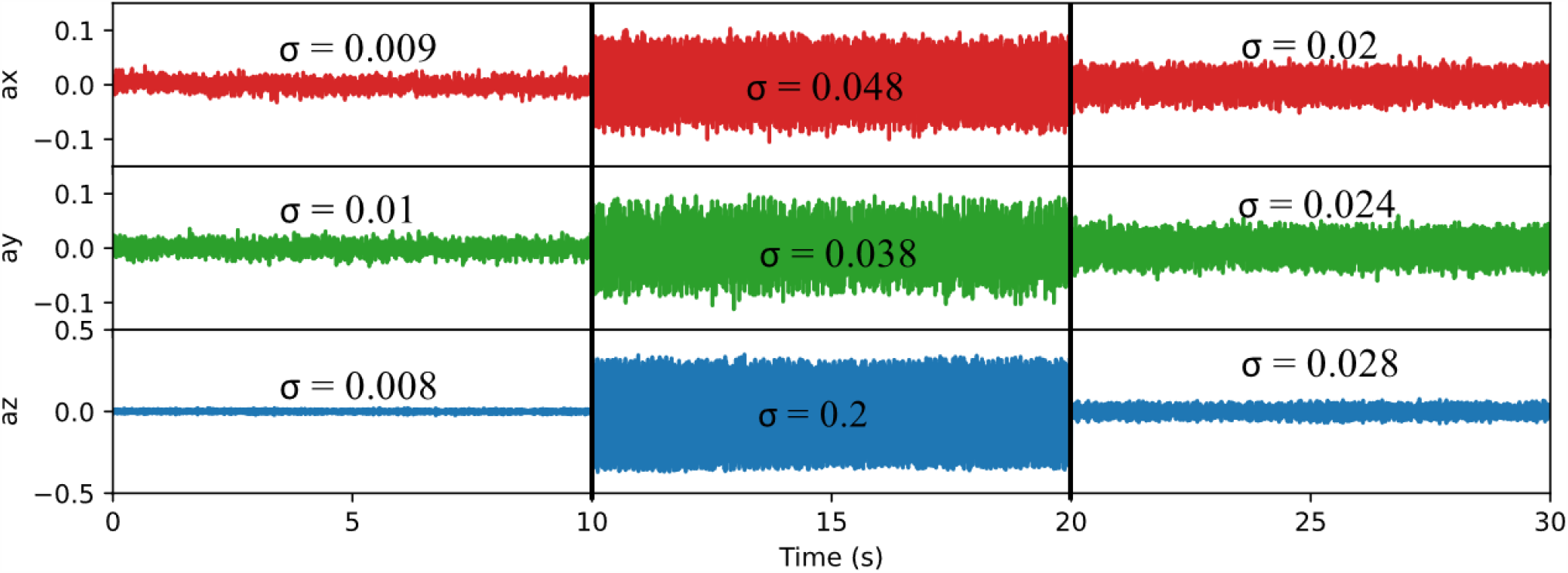
Time variation of accelerometer sensor data from a smartphone placed on the same surface next to the Mobilefuge is plotted to show the vibrations caused by the Mobilefuge. The standard deviation “σ” of the variation in accelerometer data is shown above for each data set.

To decrease these vibrations, we attached a commonly available self-adhesive damping pad at the bottom of the Mobilefuge and showed the corresponding accelerometer data in the right-side panel (20-30 s) of Fig. 4. It is clear that the damping pad improves the stability of the Mobilefuge as expected as these pads are made for damping the vibrations of household electric utilities. We also noticed that with the use of the damping pad, the noise recorded in the microphone decreased by 13dB indicating the improved stable operation of the Mobilefuge.

## Discussions

So far, we evaluated that Mobilefuge can generate enough force to separate the viral RNA from the saliva samples for further processing of samples in LAMP assay protocols. In the following, we will explain why Mobilefuge can be a better solution compared to the existing centrifuge designs for LAMP assay protocols.

### Affordable

Except the 3D printed parts, the Mobilefuge requires only two parts, i.e. a USB fan motor and a USB female to male cable. Each of these costs less than a dollar which makes the total cost of Mobilefuge extremely low. This is possible as the required speed of the centrifuge is low for the LAMP assay which can be achieved with such a small motor in USB fans.

### Simple

The Mobilefuge design presented here consists of a minimum amount of parts required for a centrifuge without sacrificing safety features. Moreover, Mobilefuge exploits the ubiquitously available USB cables for connecting the motor to a power source. Therefore, unlike other electricity powered centrifuges [29], [30], [31], [32], [33], [34], [35], [36], Mobilefuge requires neither soldering of any electrical components nor programming of microcontrollers. Thus, building a Mobilefuge device is a quick and relatively easy process even for non-experts (see Video V1). Also, there is no other equipment required other than the 3D printer to build the Mobilefuge. This simple design and build process also allows fast servicing of Mobilefuge in the case of worn or repair of individual parts which is crucial for high throughput use cases such as SARS-CoV2 testing.

### Stable

Stability of the Mobilefuge is comparatively higher due to low-speed of the device and the added damping pad which makes the Mobilefuge stable as demonstrated in Fig 4.

### Scalable

Mobilefuge can be built by anyone with a 3D-printer. During this pandemic, it has been witnessed that 3D printers are widely available to the public even in low income countries and they are willing to help to make medical equipment at home and donate to the community [20], [21], [33], [52], [53]. This is particularly important when supply chains are constrained, and demand is high for medical equipment. Therefore, if required in high volume, Mobilefuge devices can be built and distributed with the help of the 3D printing community. The low number of electric parts and their low-cost, ubiquitous availability helps to scale-up the production of the Mobilefuge. Furthermore, the size of the entire Mobilefuge is extremely compact when compared to many 3D printed centrifuge designs. Therefore, Mobilefuge’s small size allows it to be printed on even low-cost 3D printers with small print volumes such as Monoprice select Mini V2, while large 3D printers can allow to print more than one Mobilefuge in a single run. Furthermore, Mobilefuge offers easier adoption of injection moulding process for large scale manufacturing as its jar and lid designs are commodity parts that are already produced in high volume using the injection moulding.

### Low-power consumption

One of the main features of the Mobilefuge is that it can be powered from mobile phones and associated power supplies such as battery banks that can supply 5V. This is because the required rotational speed of the centrifuge is low for LAMP assays, thus Mobilefuge can be powered with such low-power supplying sources. This compelling feature makes Mobilefuge to be employed in remote and resource limited regions where the direct electric power supply is scarce and unpredictable.

### Safe

Safe operation of the equipment is crucial to convince the stakeholders to employ Mobilefuge for its intended use of detecting SARS-CoV2 near the patients in resource limited settings. Centrifuges, particularly those that have high speed rotors are notoriously unsafe [54]. Also, centrifuge tubes can be escaped from the rotors that can cause serious injuries and the leaked contents can cause biohazard. These risks are mitigated in the Mobilefuge by choosing a low-speed motor that provides just enough G-force. Additionally, screw type top lid helps to protect in case if centrifuge tubes or its contents escape from the rotor. Furthermore, use of less power hungry and consumer grade electrical components decreases the electrical hazards.

### Flexible

All the 3D-printed parts of the Mobilefuge are designed using a opensource CAD (OpenScad) software and the source files as well as STL of these parts are made freely available. Therefore, it is possible for others to optimise the design of the Mobilefuge for further improvements such as to print the entire Mobilefuge body i.e. bottom, central part and the top lid in one print or to increase the number of slots in the rotor to hold more tubes. Coming to the non 3D-printed parts of the Mobilefuge, a variety of low-power DC motors in consumer electronics devices such as electric toys, electric toothbrushes and trimmers can be used instead of the fan motor used here. Similarly, many different types of 5V power sources and the corresponding USB cables can be used to power the Mobilefuge, thus giving the flexibility to tweak according to the availability of the resources.

Additionally, one can also use a USB fan speed controller (see Fig. S1) to adjust the speed of the Mobilefuge if such features are required. The USB speed controller shown in Fig. S1 (a) allows to step up the standard 5V output from USB ports to 12V continuously, without requiring any soldering of parts. Such a step up of the voltage helps to achieve higher RPMs which will be quite useful for applications like separation of plasma from blood to diagnose malaria [24].

Overall, Mobilefuge is a simple yet highly useful equipment that can help in detection of SARS-CoV2 in saliva samples with LAMP assays. The main goal of Mobilefuge and similar efforts by other researchers is to provide accessible tools that can aid lowering the costs of SARS-CoV2 testing to contain the spread of COVID-19. Particularly, with the advent of distributed manufacturing processes, tools like Mobilefuge provide alternatives to traditional high-cost, supply chain constrained equipment.

Although the Mobilefuge is specifically aimed to be useful for LAMP assay process to detect SARS-CoV2, it is also useful for applications such as urine [33] and sperm analysis [55] and other biochemical process analysis [39], [56] where a centrifuge with low RPM/RCF is sufficient [57]. It will also serve as a useful tool to be built by school children for their science labs as it requires only a few non-3D printable parts and does not require soldering and other equipment that might not be suitable for school settings.

## Conclusions

We presented an open-source design of a 3D-printed centrifuge that is affordable, simple, stable, safe, robust, less power hungry, thus can be powered even from mobile phones. We evaluated the performance of Mobilefuge and found that it can achieve the speed of 7500 RPM and RCF of 2835, which are enough for the purification process in LAMP assay protocols. Therefore, Mobilefuge can be used to improve the sensitivity of low-cost LAMP assays, consequently making detection of SARS-CoV2 possible in saliva samples. We believe that Mobilefuge will help to lower the costs of testing of SARS-CoV2 and aid the efforts in the fight against COVID-19.

## Data Availability

All the design files are available at https://github.com/MohiniBhupathi/Mobilefuge

https://github.com/MohiniBhupathi/Mobilefuge

## Declaration of Competing Interest

The authors declare that they have no known competing financial interests or personal relationships that could have appeared to influence the work reported in this article.

## Acknowledgments

This research did not receive any specific grant from funding agencies in the public, commercial, or not-for-profit sectors. G.C.R.D. acknowledges funding from the European Union’s Horizon2020 research and innovation programme under Marie Sklodowska-Curie grant agreement No. 713654. M.T for his time during this project.

## Supplementary material

All the design files are available at https://github.com/MohiniBhupathi/Mobilefuge

## Notes

### Competing Interest Statement

The authors have declared no competing interest.

### Author Declarations

There are no humans or animals invloved in this study, so I don't think any of these fields in this sectin are relevant. But for the sake of getting the article past this barrier, I am consenting of all these approvals.

## References

[1] D. Baud, X. Qi, K. Nielsen-Saines, D. Musso, L. Pomar, G. Favre, Real estimates of mortality following COVID-19 infection, Lancet Infect. Dis. 20 (2020) 773.

[2] M. Nicola, Z. Alsafi, C. Sohrabi, A. Kerwan, A. Al-Jabir, C. Iosifidis, M. Agha, R. Agha, The socio-economic implications of the coronavirus pandemic (COVID-19): A review, Int. J. Surg. 78 (2020) 185–193.

[3] Molecular and antibody point-of-care tests to support the screening, diagnosis and monitoring of COVID-19 - CEBM, CEBM. (2020). https://www.cebm.net/covid-19/molecular-and-antibody-point-of-care-tests-to-support-the-screening-diagnosis-and-monitoring-of-covid-19 / (accessed July 15, 2020).

[4] R. Weissleder, H. Lee, J. Ko, M.J. Pittet, COVID-19 diagnostics in context, Sci. Transl. Med. 12 (2020). https://doi.org/10.1126/scitranslmed.abc1931.

[5] J. Kashir, A. Yaqinuddin, Loop mediated isothermal amplification (LAMP) assays as a rapid diagnostic for COVID-19, Med. Hypotheses. 141 (2020) 109786.

[6] M.J. Kellner, J.J. Ross, J. Schnabl, M.P.S. Dekens, R. Heinen, N.A. Tanner, R. Fritsche-Polanz, M. Traugott, T. Seitz, A. Zoufaly, M. Födinger, C. Wenisch, J. Zuber, Vienna Covid-19 Diagnostics Initiative (VCDI), A. Pauli, J. Brennecke, Scalable, rapid and highly sensitive isothermal detection of SARS-CoV-2 for laboratory and home testing, Cold Spring Harbor Laboratory. (2020) 2020.06.23.166397. https://doi.org/10.1101/2020.06.23.166397.

[7] SnapDx, (2020). https://www.snapdx.org (accessed December 27, 2020).

[8] Emergency use authorization (EUA) summary for the color SARS-CoV-2 RT-LAMP diagnostic assay, (2020). https://www.fda.gov/media/138249/download (accessed July 23, 2020).

[9] Office of the Commissioner, Coronavirus (COVID-19) Update: FDA Authorizes First Diagnostic Test Using At-Home Collection of Saliva Specimens, U.S. Food and Drug Administration. (2020). https://www.fda.gov/news-events/press-announcements/coronavirus-covid-19-update-fda-authorizes-first-diagnostic-test-using-home-collection-saliva (accessed July 15, 2020).

[10] AQ-TOP ™ COVID-19 Rapid Detection Kit, (2020). https://www.fda.gov/media/138307/download (accessed July 23, 2020).

[11] E. Li, A. Larson, A. Kothari, M. Prakash, Handyfuge-LAMP: low-cost and electricity-free centrifugation for isothermal SARS-CoV-2 detection in saliva, (2020). https://doi.org/10.1101/2020.06.30.20143255.

[12] B.A. Rabe, C. Cepko, SARS-CoV-2 Detection Using an Isothermal Amplification Reaction and a Rapid, Inexpensive Protocol for Sample Inactivation and Purification, (2020). https://doi.org/10.1101/2020.04.23.20076877.

[13] M.J. Kellner, J.J. Ross, J. Schnabl, M.P.S. Dekens, R. Heinen, I. Grishkovskaya, B. Bauer, J. Stadlmann, L. Menéndez-Arias, R. Fritsche-Polanz, M. Traugott, T. Seitz, A. Zoufaly, M. Födinger, C. Wenisch, J. Zuber, Vienna Covid-19 Diagnostics Initiative (VCDI), A. Pauli, J. Brennecke, A rapid, highly sensitive and open-access SARS-CoV-2 detection assay for laboratory and home testing, bioRxiv. (2020) 2020.06.23.166397. https://doi.org/10.1101/2020.06.23.166397.

[14] V.L.D. Thi, K. Herbst, K. Boerner, M. Meurer, L.P.M. Kremer, D. Kirrmaier, A. Freistaedter, D. Papagiannidis, C. Galmozzi, M.L. Stanifer, S. Boulant, S. Klein, P. Chlanda, D. Khalid, I.B. Miranda, P. Schnitzler, H.-G. Kräusslich, M. Knop, S. Anders, A colorimetric RT-LAMP assay and LAMP-sequencing for detecting SARS-CoV-2 RNA in clinical samples, Sci. Transl. Med. (2020). https://doi.org/10.1126/scitranslmed.abc7075.

[15] One Hour Covid Test using LAMP, (2020). https://app.jogl.io/project/163/1HourCovidTest (accessed January 2, 2021).

[16] M. El-Tholoth, H.H. Bau, J. Song, A Single and Two-Stage, Closed-Tube, Molecular Test for the 2019 Novel Coronavirus (COVID-19) at Home, Clinic, and Points of Entry, ChemRxiv. (2020). https://doi.org/10.26434/chemrxiv.11860137.

[17] D. Guan, D. Wang, S. Hallegatte, S.J. Davis, J. Huo, S. Li, Y. Bai, T. Lei, Q. Xue, D.’maris Coffman, D. Cheng, P. Chen, X. Liang, B. Xu, X. Lu, S. Wang, K. Hubacek, P. Gong, Global supply-chain effects of COVID-19 control measures, Nat Hum Behav. 4 (2020) 577–587.

[18] I. Boškoski, C. Gallo, M.B. Wallace, G. Costamagna, COVID-19 pandemic and personal protective equipment shortage: protective efficacy comparing masks and scientific methods for respirator reuse, Gastrointest. Endosc. (2020). https://doi.org/10.1016/j.gie.2020.04.048.

[19] M.L. Ranney, V. Griffeth, A.K. Jha, Critical Supply Shortages — The Need for Ventilators and Personal Protective Equipment during the Covid-19 Pandemic, New England Journal of Medicine. 382 (2020) e41. https://doi.org/10.1056/nejmp2006141.

[20] J.M. Pearce, Distributed Manufacturing of Open Source Medical Hardware for Pandemics, J. Mater. Process. Manuf. Sci. 4 (2020) 49.

[21] M. Javaid, A. Haleem, R. Vaishya, S. Bahl, R. Suman, A. Vaish, Industry 4.0 technologies and their applications in fighting COVID-19 pandemic, Diabetes Metab. Syndr. 14 (2020) 419–422.

[22] C.-H. Liu, C.-A. Chen, S.-J. Chen, T.-T. Tsai, C.-C. Chu, C.-C. Chang, C.-F. Chen, Blood Plasma Separation Using a Fidget-Spinner, Anal. Chem. 91 (2019) 1247–1253.

[23] I. Michael, D. Kim, O. Gulenko, S. Kumar, S. Kumar, J. Clara, D.Y. Ki, J. Park, H.Y. Jeong, T.S. Kim, S. Kwon, Y.-K. Cho, A fidget spinner for the point-of-care diagnosis of urinary tract infection, Nat Biomed Eng. 4 (2020) 591–600.

[24] M.S. Bhamla, M. Saad Bhamla, B. Benson, C. Chai, G. Katsikis, A. Johri, M. Prakash, Paperfuge: An ultra-low cost, hand-powered centrifuge inspired by the mechanics of a whirligig toy, (2020). https://doi.org/10.1101/072207.

[25] G. Byagathvalli, A. Pomerantz, S. Sinha, J. Standeven, M. Saad Bhamla, A 3D-printed hand-powered centrifuge for molecular biology: Supplementary Information, (2016). https://doi.org/10.1101/519835.

[26] J. Brown, L. Theis, K. O’Connor, L. Kerr, M. Uthman, Z. Maria Oden, N. Zakhidova, R. Richards-Kortum, A Hand-Powered, Portable, Low-Cost Centrifuge for Diagnosing Anemia in Low-Resource Settings, The American Journal of Tropical Medicine and Hygiene. 85 (2011) 327–332. https://doi.org/10.4269/ajtmh.2011.10-0399.

[27] Science Buddies, Spin Zone: Separating Butter with a Salad Spinner Centrifuge | Science Project, (2012). https://www.sciencebuddies.org/science-fair-projects/project-ideas/BioChem_p040/biotechnology-techniques/separating-butter-with-a-salad-spinner-centrifuge (accessed July 16, 2020).

[28] A.P. Wong, M. Gupta, S.S. Shevkoplyas, G.M. Whitesides, Egg beater as centrifuge: isolating human blood plasma from whole blood in resource-poor settings, Lab on a Chip. 8 (2008) 2032. https://doi.org/10.1039/b809830c.

[29] One-Piece Centrifuge for Rotary Tools, Thingiverse. (2020). https://www.thingiverse.com/thing:1483 (accessed July 14, 2020).

[30] A. Priye, S. Wong, Y. Bi, M. Carpio, J. Chang, M. Coen, D. Cope, J. Harris, J. Johnson, A. Keller, R. Lim, S. Lu, A. Millard, A. Pangelinan, N. Patel, L. Smith, K. Chan, V.M. Ugaz, Lab-on-a-Drone: Toward Pinpoint Deployment of Smartphone-Enabled Nucleic Acid-Based Diagnostics for Mobile Health Care, Anal. Chem. 88 (2016) 4651–4660.

[31] F. Lab’s DIYbio Centrifuge, Thingiverse.com. (2015). https://www.thingiverse.com/thing:1175393 (accessed July 14, 2020).

[32] BioHackAcademy/BHA_Centrifuge, GitHub. (2016). https://github.com/BioHackAcademy/BHA_Centrifuge (accessed July 15, 2020).

[33] Behance, The Frugal Centrifuge, Behance. (2019). https://www.behance.net/gallery/86450155/The-Frugal-Centrifuge (accessed July 16, 2020).

[34] Polyfuge: A DIY Open-Source Microcentrifuge for Everyone, Kickstarter. (2018). https://www.kickstarter.com/projects/1733191226/polyfuge-a-diy-open-source-microcentrifuge-for-eve (accessed July 16, 2020).

[35] Instructables, OpenFuge, (2013). https://www.instructables.com/id/OpenFuge / (accessed July 16, 2020).

[36] Centrifuge, Bento Lab. (2015). https://www.bento.bio/resources/manual/centrifuge / (accessed July 29, 2020).

[37] Z. WareJoncas, C. Stewart, J. Giannini, An Inexpensive, Open-Source Mini-Centrifuge, Am. Biol. Teach. 80 (2018) 451–456.

[38] Hacked Hard-disc Centrifuge - Hackteria Wiki, (2016). https://hackteria.org/wiki/Hacked_Hard-disc_Centrifuge (accessed July 14, 2020).

[39] H. Ramachandraiah, M. Amasia, J. Cole, P. Sheard, S. Pickhaver, C. Walker, V. Wirta, P. Lexow, R. Lione, A. Russom, Lab-on-DVD: standard DVD drives as a novel laser scanning microscope for image based point of care diagnostics, Lab Chip. 13 (2013) 1578–1585.

[40] DremelFuge - A One-Piece Centrifuge for Rotary Tools, Thingiverse. (2009). https://www.thingiverse.com/thing:1483 (accessed July 14, 2020).

[41] USB On-The-Go and Embedded Host, Web.archive. (2011). https://web.archive.org/web/20120505091034/http://www.usb.org/developers/onthego/ (accessed December 27, 2020).

[42] OpenSCAD, (2019). http://openscad.org (accessed July 14, 2020).

[43] Customizable Container with Knurled Lid, Thingiverse.com. (2014). https://www.thingiverse.com/thing:612709 (accessed July 14, 2020).

[44] Parametric airless tire, Thingiverse. (2012). https://www.thingiverse.com/thing:17514 (accessed July 14, 2020).

[45] Microfuge, Github. (2020). https://github.com/MohiniBhupathi/Microfuge (accessed December 28, 2020).

[46] Monoprice select mini V2, Monoprice. (2020). https://www.monoprice.com/product?p_id=15365 (accessed July 16, 2020).

[47] D. Halliday, R. Resnick, J. Walker, Fundamentals of Physics, Part 2, Chapters 13 - 21, John Wiley & Sons Incorporated, 2000.

[48] D. Halliday, R. Resnick, G.H. Bowen, Fundamentals of Physics, Physics Today. 25 (1972) 53–54. https://doi.org/10.1063/1.3070817.

[49] B. Vogelstein, D. Gillespie, Preparative and analytical purification of DNA from agarose, Proceedings of the National Academy of Sciences. 76 (1979) 615–619. https://doi.org/10.1073/pnas.76.2.615.

[50] R. Boom, C.J. Sol, M.M. Salimans, C.L. Jansen, P.M. Wertheim-van Dillen, J. van der Noordaa, Rapid and simple method for purification of nucleic acids, Journal of Clinical Microbiology. 28 (1990) 495–503. https://doi.org/10.1128/jcm.28.3.495-503.1990.

[51] S. Staacks, Your smartphone is a mobile lab, Phyphox. (2016). https://phyphox.org (accessed August 29, 2020).

[52] COVID19-APHP Group. Electronic address, Assistance Publique-Hôpitaux de Paris’ response to the COVID-19 pandemic, Lancet. 395 (2020) 1760–1761.

[53] S. Singh, C. Prakash, S. Ramakrishna, Three-dimensional printing in the fight against novel virus COVID-19: Technology helping society during an infectious disease pandemic, Technol. Soc. 62 (2020) 101305.

[54] D.E. Clark, Safety and the laboratory centrifuge, Chemical Health and Safety. 8 (2001) 7–13. https://doi.org/10.1016/s1074-9098(01)00263-5.

[55] Trak® Fertility - Top Rated At Home Male Fertility Testing System, Trak Fertility. (2020). https://trakfertility.com/ (accessed July 16, 2020).

[56] I.J. Michael, T.-H. Kim, V. Sunkara, Y.-K. Cho, Challenges and Opportunities of Centrifugal Microfluidics for Extreme Point-of-Care Testing, Micromachines (Basel). 7 (2016). https://doi.org/10.3390/mi7020032.

[57] D. Rickwood, T. Ford, J. Steensgaard, Centrifugation: Essential Data, John Wiley & Son Limited, 1994.

