## Supplementary material for "Mobilefuge: A low-cost, portable, open source, 3D-printed centrifuge that can be used for purification of saliva samples for SARS-CoV2 detection": Figure S1

1. Rossa Lodge, Rossa Avenue, Bishopstown, Cork, T12YWR0, Ireland.

2. Centre for Advanced Photonics and Process Analysis, Cork Institute of Technology, Cork, T12P928, Ireland.

3. Tyndall National Institute, Cork, T12R5C, Ireland

### Abstract:

One of the best ways to contain the spread of COVID-19 is frequent testing of as many people as possible and timely isolation of uninfected personnel from infected personnel. However, the cost of massive testing is affordable in many countries. The existing technologies might not be scalable to offer affordable testing for millions of people. To address this issue, novel testing methods based on Loop-Mediated Isothermal Amplification (LAMP) were proposed that are more sensitive, require less reagents and can work with saliva samples instead of more tedious nasal swabs. As a result, LAMP based protocols can make it possible to drive the cost down to one dollar per test. These LAMP based methods require a centrifuge device, mostly for separation of viral particles from reaction inhibitors in saliva samples. However, centrifuge is neither accessible nor affordable in many resource limited settings, especially during this pandemic situation when normal supply chains are heavily disrupted. To overcome these challenges, we invented a low-cost centrifuge that can be useful for carrying out low-cost LAMP based detection of SARS-Cov2 virus in saliva. The 3D printed centrifuge (Mobilefuge) is portable, robust, stable, safe, easy to build and operate. The Mobilefuge doesn't require soldering or programming skills and can be built without any specialised equipment, yet practical enough for high throughput use. More importantly, Mobilefuge can be powered from widely available USB ports, including mobile phones and associated power supplies. This allows the Mobilefuge to be used even in off-grid and resource limited settings. We believe that our invention will aid the efforts to contain the spread of COVID-19 by lowering the costs of testing equipment. Apart from the COVID-19 testing, the Mobilefuge can have applications in the field of biomedical research and diagnostics.

### Supplementary material

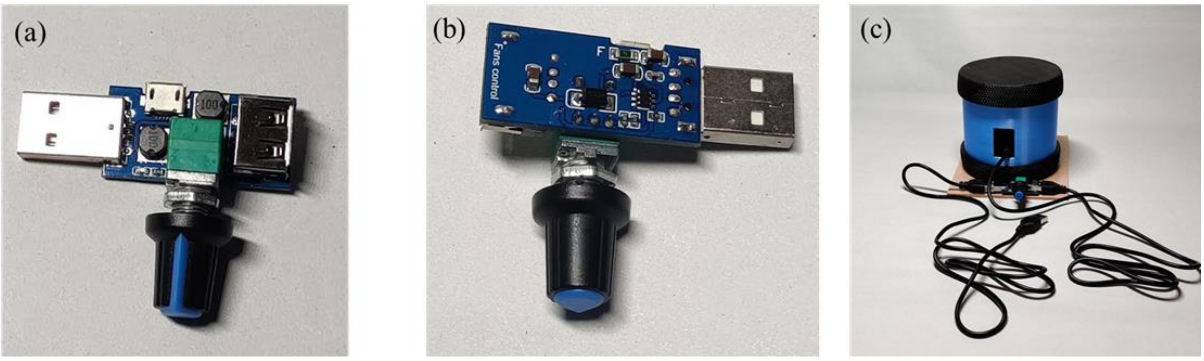


Figure S1: (a) Front view of the USB fan speed controller (b) Back view of the USB speed controller (c) Mobilefuge with USB fan controller to control the speed of the Mobilefuge.
